# A metabolite-based machine learning approach to diagnose Alzheimer’s-type dementia in blood: Results from the European Medical Information Framework for Alzheimer’s Disease biomarker discovery cohort

**DOI:** 10.1101/19007146

**Authors:** Daniel Stamate, Min Kim, Petroula Proitsi, Sarah Westwood, Alison Baird, Alejo Nevado-Holgado, Abdul Hye, Isabelle Bos, Stephanie Vos, Rik Vandenberghe, Charlotte E. Teunissen, Mara Ten Kate, Philip Scheltens, Silvy Gabel, Karen Meersmans, Olivier Blin, Jill Richardson, Ellen De Roeck, Sebastiaan Engelborghs, Kristel Sleegers, Régis Bordet, Lorena Rami, Petronella Kettunen, Magda Tsolaki, Frans Verhey, Daniel Alcolea, Alberto Lléo, Gwendoline Peyratout, Mikel Tainta, Peter Johannsen, Yvonne Freund-Levi, Lutz Frölich, Valerija Dobricic, Giovanni B Frisoni, José L Molinuevo, Anders Wallin, Julius Popp, Pablo Martinez-Lage, Lars Bertram, Kaj Blennow, Henrik Zetterberg, Johannes Streffer, Pieter J Visser, Simon Lovestone, Cristina Legido-Quigley

## Abstract

**INTRODUCTION:** Machine learning (ML) may harbor the potential to capture the metabolic complexity in Alzheimer’s Disease (AD). Here we set out to test the performance of metabolites in blood to categorise AD when compared to CSF biomarkers.

**METHODS:** This study analysed samples from 242 cognitively normal (CN) people and 115 with AD-type dementia utilizing plasma metabolites (n=883). Deep Learning (DL), Extreme Gradient Boosting (XGBoost) and Random Forest (RF) were used to differentiate AD from CN. These models were internally validated using Nested Cross Validation (NCV).

**RESULTS:** On the test data, DL produced the AUC of 0.85 (0.80-0.89), XGBoost produced 0.88 (0.86-0.89) and RF produced 0.85 (0.83-0.87). By comparison, CSF measures of amyloid, p-tau and t-tau (together with age and gender) produced with XGBoost the AUC values of 0.78, 0.83 and 0.87, respectively.

**DISCUSSION:** This study showed that plasma metabolites have the potential to match the AUC of well-established AD CSF biomarkers in a relatively small cohort. Further studies in independent cohorts are needed to validate whether this specific panel of blood metabolites can separate AD from controls, and how specific it is for AD as compared with other neurodegenerative disorders

## 1. Introduction

At present, the diagnosis of Alzheimer’s disease–type dementia (AD) is based on protein biomarkers in cerebrospinal fluid (CSF) and brain imaging together with a battery of cognition tests. Diagnostic tools based on CSF collection are invasive while brain-imaging tools are still costly, and therefore, there is a need to identify non-invasive tools for early detection as well as for measuring disease progression.

In recent years, an increasing number of studies have examined blood metabolites as potential AD biomarkers [1-4]. The advantages of looking at blood metabolites are that they are easily accessible but also that they represent an essential aspect of the phenotype of an organism and hence might act as a molecular fingerprint of disease progression [5, 6]. Therefore, blood AD markers could potentially aid early diagnosis and recruitment for trials.

Here we utilised data generated as part of the European Medical Information Framework for AD Multimodal Biomarker Discovery (EMIF-AD) previously reported in full in Kim et al [8]. As discussed in that paper, metabolite levels were measured using liquid chromatography–mass spectroscopy (LC-MS) to cover ca. 800 metabolites and these metabolites related to CSF biomarkers of AD commonly used in clinical research including trials, and increasingly in clinical practice, as part of the diagnostic work up. Here we explore the potential of different machine Learning (ML) algorithms to identify those individuals with AD from dataset and to compare the effectiveness of blood based metabolites as an indicator of clinical diagnosis to that of CSF markers. In this study we employed two state of the art ML algorithms - Deep Learning (DL) and Extreme Gradient Boosting (XGBoost) - and compared these to the more commonly utilized Random Forest (RF) algorithm.

## 2. Methods

This study accessed data previously generated from 242 samples from cognitively normal (CN) individuals and 115 from people with AD-type dementia (AD) samples in which diagnosis was based on clinical diagnosis. Details on the subjects, clinical and cognitive data as well as measurements of AD pathological markers have been described elsewhere [7, 8]. The metabolomics data employed here was accessed in the EMIF-AD portal and the acquisition and processing details can be found via open access in [8]. In short, the EMIF-AD cohort is a collated cohort making use of existing data and samples collected in 11 different studies across Europe, with the aim to discover novel diagnostic and prognostic markers for predementia AD.

In the current study, the main objective was to use state of the art ML classification algorithms to build CN vs AD predictive models using blood metabolites. For this purpose, we employed DL and XGBoost. Additionally we also employed the more popularly used RF) algorithm. These models were compared in terms of binary classifiers with Area Under the Curve (AUC) in Receiver Operating Characteristic (ROC) curves.

The metabolites with more than 45% missing values were discarded. The remaining missing values were handled with imputation methods based on the k-nearest neighbour (RF and DL), or internally by the classification algorithm (XGBoost). Models were built and evaluated using a Nested Cross Validation (NCV) which used 9/10 data folds for model training and optimisation in an inner cross validation, and 1/10 data folds for model testing in an outer cross validation. The process was repeated 10 times, for each of the test data folds.

The analysis was further extended by assessing the stability of the AUC performance with Monte Carlo (MC) simulations consisting of 50 repeated similar NCV experiments. As such, multiple models were built on multiple samples in the NCV and MC, using metabolite predictors selected on the basis of their capability to discriminate CN vs AD as measured by the Relief algorithm [9] applied on training data in combination with 500 permutations of the outcome variable’s values. This method computes the predictors’ importance defined as the standardised Relief score, according to Measuring Predictor Importance chapter of [10]. Part of the prediction modelling methodology in this study was adapted after [11], with different algorithms, and followed recommendations from [10, 12]. The analysis was carried out using R software [13]. Pathway analysis was performed on the top 20 ranked metabolites using MetaboAnalyst 4.0 [14]. The algorithms were run on four servers with 6-core Xeon CPUs and 336 GB RAM.

## 3. Results

In this study, we analysed metabolite data derived from blood samples from 358 participants (CN n=242, AD n=115) previously reported in Kim et al [8]. Demographic and clinical data can be found in [8]; in short, there was no difference in gender while AD participants were older when compared with CN participants.

On the test data, the DL model produced a Receiver Operating Characteristic (ROC) Area Under the Curve (AUC) value of 0.85 with its 95% confidence interval (CI) ranging between [0.8038, 0.8895]. The XGBoost model produced the AUC value of 0.88 (95% CI [0.8619, 0.8903]). When the classifier model RF was employed, the resulting AUC was 0.85 (95%CI [0.8323, 0.8659]). Fig. 1 illustrates ROC curves obtained from the three ML models.

**Figure 1.**
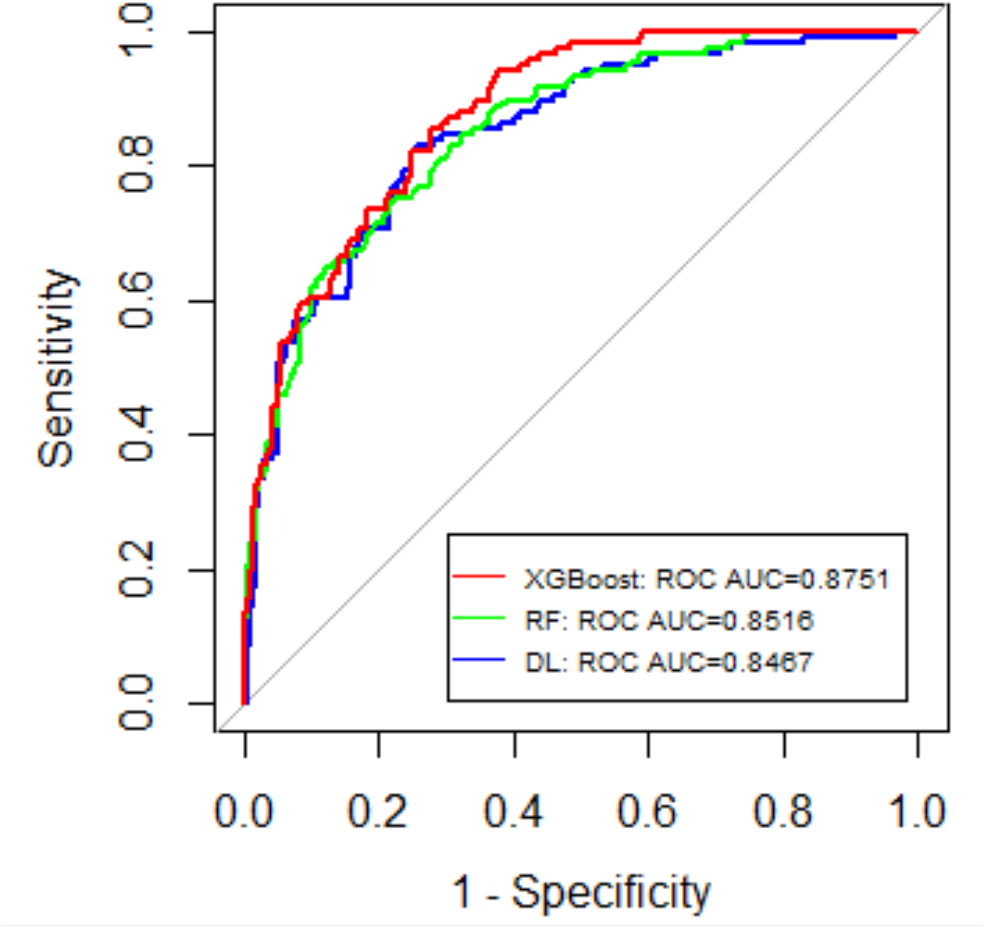
Shows the AUC values for the XGBoost, RF and DL models. XGBoost performed best with metabolite predictors in the EMIF cohort.

The MC simulation conducted with XGBoost which was the superior predictive model in our analysis, led to a Gaussian distribution of the AUC values according to [11] and as confirmed by Shapiro-Wilk test (p-value=0.6819). The 50 AUC values obtained in MC had a minimum of 0.8614, a maximum of 0.8923, a mean of 0.8761, a median of 0.8766, and a standard deviation of 0.0072. The t-test showed that the true mean of AUC for XGBoost applied on plasma metabolites was not lower than 0.87 (p-value=1.265×10^−07^).

For comparison, we also investigated the levels of amyloid, p-tau and t-tau, to which we added also age and gender, and their prediction for clinical AD vs CN. XGBoost models were built in the same manner as for metabolite predictors. Together with age and gender, amyloid led to AUC 0.78 (95%CI [0.7626, 0.8013]); p-tau led to AUC 0.83 (95%CI [0.8188, 0.8470]); and t-tau led to AUC 0.87 (95%CI [0.8583, 0.8854]). From the mean AUC for metabolites and for amyloid, p-tau and t-tau calculated individually, the t-tests showed superior values for metabolites (p-value<2.2×10^−16^, p-value<2.2×10^−16^, and p-value=0.005921, respectively).

The top 20 ranked predictors out of the 347 selected by the method presented in the previous section, are shown in Fig. 2.

**Figure 2.**
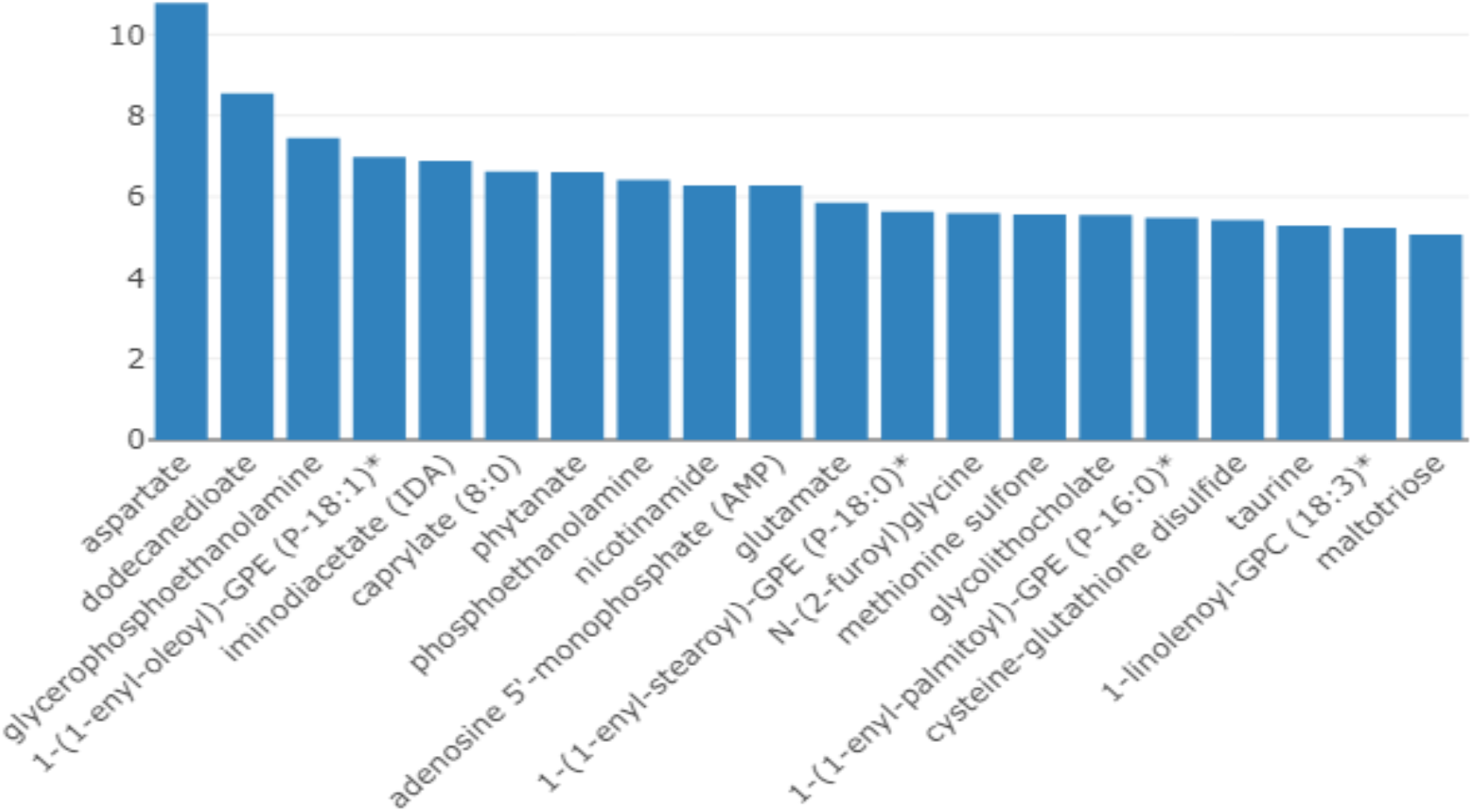
The x-axis shows the top 20 ranked predictors, and the *y*-axis shows the predictors’ importance computed as the standardised Relief score according to Measuring Predictor Importance chapter of [10].

Pathway analyses revealed that the Nitrogen pathway was overrepresented (q_FDR_=0.004) within the panel. Molecules that were captured as the 20 top ranking predictors are discussed in the next section.

## 4. Discussion

Machine Learning applied to healthcare is increasingly enabled by the advent of high-performance computing and the development of complex algorithms. In this study, we employed two state of the art algorithms, DL and XGBoost, and a more conventional algorithm, RF, to obtain high accuracy models to predict AD vs CN with metabolites as predictors. Our study showed that the best model was based on XGBoost [15] which is an enhanced form of Gradient Boosting Machines methods based on decision trees [12]. In our study RF and DL achieved comparable AUC. DL algorithms are known to often take advantage of large and/or unstructured data (such as images) to produce more accurate category discrimination/ prediction. In a study using the Alzheimer’s Disease Neuroimaging Initiative (ADNI) data for AD prediction, XGBoost demonstrated superior results (AUC= 0.97 (0.01)) when including imaging parameters (MRI and PET) as predictors and when compared to RF, Support Vector Machines, Gaussian Processes and Stochastic Gradient Boosting [16]. In other study where cognition and MRI were used as predictors, Kernel Ridge Regression performed to R^2^=0.87 (0.025) when cognition and MRI predictors were included [17].

Pathway analyses using the top 20 AD predicting metabolites derived from the Relief method showed that the Nitrogen pathway was overrepresented. Some of the molecules selected have been reported in metabolomics studies and have been implicated in neurodegeneration: Dodecanate, which is a C12 fatty acid, was found correlated to longitudinal measures of cognition in the ADNI cohort [3] and so was the bile acid glycolithocholate which was associated to both AD and cognition measures (ADAS-Cog13) in one of the biggest cross-sectional studies on cognition, AD and the microbiome [18]. Plasmalogens were also found in decreased levels in our cohort in agreement with an earlier report [19]. The amide form of vitamin B3, nicotamide, has been implicated in both neuroprotection and neuronal death [20].

New metabolites that could be of interest and have not been previously reported as related to AD were phytanate and furoylglycine. The former is a known neurotoxin which impairs mitochondrial function and transcription [21]. Furoylglycine is a metabolite which, as lithocholic acid, is mainly synthesized by the microbiome and has been reported as a biomarker of coffee consumption [22].

A limitation of our study is that it does not include an external validation due to the size of the cohort. However, we implemented a NCV procedure repeated 50 times in a MC simulation that led to an extended internal validation with prediction accuracy of cases. Further studies will assess the performance of ratios/combinations of CSF markers and metabolites, life-style factors and disorders commonly found in the elderly, together with testing the specificity for this specific panel in other neurodegenerative (e.g. PD, FTD), neurological (e.g. stroke) and psychiatric (e.g. depression) disorders associated with aging.

The intent of this paper was to compare the performance of different ML algorithms to identify people with AD from cognitively unimpaired individuals. Here we show first that all three approaches used demonstrate good discriminatory power, second that XGBoost is somewhat more effective in this particular dataset than RF and DL and third, that this accuracy for clinical diagnosis is broadly similar to that achieved by CSF markers of AD pathology. The lack of a replication and validation dataset limits the interpretation of this finding but nonetheless the strong prediction of diagnostic category from a blood based metabolite biomarker set is further evidence of the potential of such approaches to complement other biomarkers in identification of people with likely AD

## Data Availability

On request to the data repository’s

## 6. Acknowledgments

The authors thank the individuals and families who took part in this research. The authors would also like to thank all people involved in data and sample collection and/or logistics across the different centers, and in particular Marije Benedictus, Wiesje van de Flier, Charlotte Teunissen, Ellen De Roeck, Naomi De Roeck, Ellis Niemantsverdriet, Charisse Somers, Babette Reijs, Andrea Izagirre Otaegi, Mirian, Ecay Torres, Sindre Rolstad, Eva Bringman, Domile Tautvydaite, Barbara Moullet, Charlotte Evenepoel, Isabelle Cleynen, Bea Bosch, Daniel Alcolea Rodriguez, Moira Marizzoni, Alberto Redolfi, and Paolo Bosco.

## Funding

The present study was conducted as part of the EMIF-AD project, which has received support from the Innovative Medicines Initiative Joint Undertaking under EMIF grant agreement no. 115372, resources of which are composed of financial contribution from the European Union’s Seventh Framework Program (FP7/2007-2013) and EFPIA companies’ in-kind contribution. The DESCRIPA study was funded by the European Commission within the fifth framework program (QLRT-2001-2455). The EDAR study was funded by the European Commission within the fifth framework program (contract no. 37670). The San Sebastian GAP study is partially funded by the Department of Health of the Basque Government (allocation 17.0.1.08.12.0000.2.454.01. 41142.001.H). Kristel Sleegers is supported by the Research Fund of the University of Antwerp. Daniel Stamate is supported by the Alzheimer’s Research UK (ARUK-PRRF2017-012).

## Notes

### Competing Interest Statement

The authors have declared no competing interest.

### Funding Statement

IMI has

### Author Declarations

All relevant ethical guidelines have been followed and any necessary IRB and/or ethics committee approvals have been obtained.

Any clinical trials involved have been registered with an ICMJE-approved registry such as ClinicalTrials.gov and the trial ID is included in the manuscript.

